# Analysis of SARS-CoV-2 transmission in different settings, among cases and close contacts from the Tablighi cluster in Brunei Darussalam

**DOI:** 10.1101/2020.05.04.20090043

**Authors:** Liling Chaw, Wee Chian Koh, Sirajul Adli Jamaludin, Lin Naing, Mohammad Fathi Alikhan, Justin Wong

## Abstract

We report the transmission dynamics of SARS-CoV-2 across different settings from the initial COVID-19 cluster in Brunei, arisen from 19 attendees at the Malaysian Tablighi Jama’at gathering and resulted in 52 locally transmitted cases. Highest non-primary attack rates(ARs) were observed at a subsequent local religious gathering (14.8% [95%CI: 7.1,27.7]) and in the household (10.6% [95%CI: 7.3,15.1]. Household ARs of symptomatic cases were higher (14.4% [95%CI: 8.8,19.9]) than asymptomatic (4.4% [95%CI: 0.0,10.5]) or presymptomatic cases (6.1% [95%CI: 0.3,11.8]). Low ARs (<1%) were observed for workplace and social settings. Our analysis highlights that SARS-CoV-2 transmission varies depending on environmental, behavioural and host factors. We identify ‘red flags’ of potential super-spreading events, namely densely populated gatherings for prolonged periods in enclosed settings, presence of individuals with recent travel history, and group behaviours such as communal eating, sleeping and sharing of personal hygiene facilities. We propose differentiated testing strategies that account for transmission risk.

**Article summary line:** We highlight the variability of SARS-CoV-2 transmission across different settings, identify settings at highest risk, and characterize the role of environmental, behavioural, and host factors in driving SARS-CoV-2 transmission.

## INTRODUCTION

Initially reported on December 31, 2019, cases of coronavirus disease (COVID-19) have since escalated significantly, prompting the World Health Organisation’s declaration of a pandemic on March 11. A rapid response by the global scientific community has described many aspects of the SARS-CoV-2 virus. Its primary mode of transmission is through respiratory droplets, and fomite and aerosol transmission may also play a role (1). Asymptomatic transmission has been observed (2) and the peak infective period appears to be within the first few days of symptom onset (3). Estimates suggest a basic reproduction number (R_0_) of 2–3 in the early stages of an outbreak (4). While the R_0_ is valuable in assessing the spread of the outbreak, it can obscure individual heterogeneity in the level of infectivity, among persons and in different settings (5, 6). Modelling of the 2003 SARS outbreak indicates that >70% of transmission occurred due to super-spreading events (SSE) (7). Early reports suggest that similar dynamics may be in play in the explosive propagation of SARS-CoV-2 (8).

As part of their mitigation strategies, many countries have adopted stringent yet blunt ‘lockdowns’ of whole cities and provinces (9) and are implementing exit strategies. In moving towards a more targeted approach, countries should be able to answer two major questions regarding the transmission dynamics of SARS-CoV-2: (i) what are the environmental, behavioural, and host factors that drive transmission? and, (ii) what are the most effective interventions to control this? To address these, we report on analysis of a transmission chain in Brunei Darussalam that resulted from an international SSE.

Brunei (population 459,500) (10) detected its first COVID-19 case on March 9, arising from an Islamic religious gathering (Tablighi Jama’at) in Kuala Lumpur, Malaysia, a known SSE lasting four days and attended by >16,000 people, including international participants (11). Of the 135 confirmed cases in Brunei reported as of first week of April, 71 cases (52.6%) have an epidemiological link to the Tablighi event (Figure 1).

**Figure 1.**
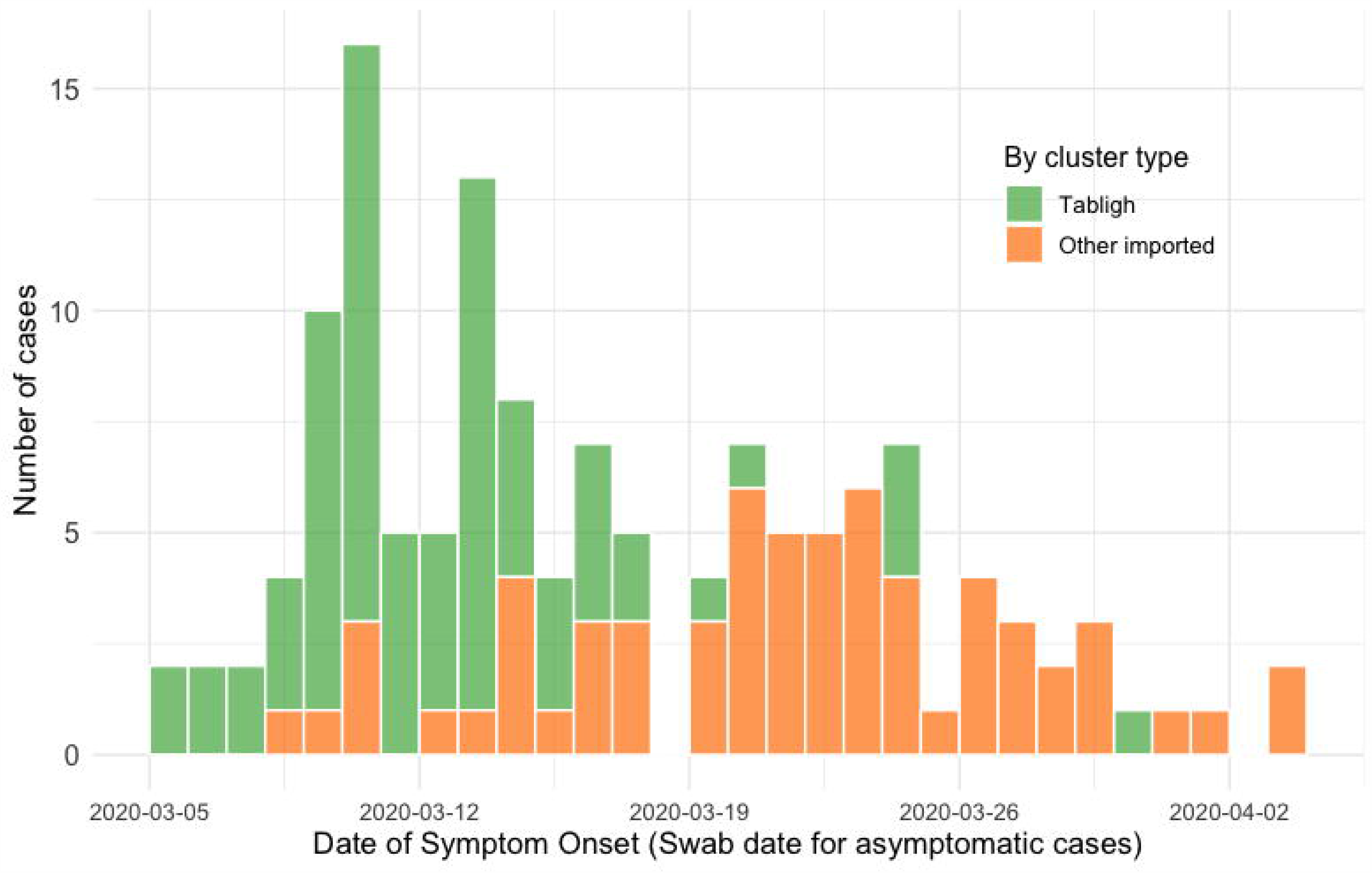
Epidemic curve for the first 135 COVID-19 cases in Brunei Darussalam, by Tablighi (green bars) and other imported (orange bars) clusters.

As SARS-CoV-2 is a novel infection in a naive population, an outbreak investigation of this event can provide insights into the transmission dynamics of the disease and the effectiveness of outbreak control measures. The thorough nature of Brunei’s contact tracing provides a rare opportunity to study the epidemiological and transmission characteristics of SARS-CoV-2 in the community setting.

## METHODS

### Surveillance and case identification

Brunei’s Ministry of Health has responsibility for communicable disease surveillance. Since January 23, testing criteria have been implemented for suspected COVID-19 cases. Initially, individuals with acute respiratory symptoms, and a travel history to a high-risk area were tested for SARS-CoV-2. Over the next weeks, the program expanded to include: (i) contacts of a confirmed case (regardless of symptoms); (ii) individuals admitted to an inpatient facility with pneumonia; and (iii) individuals who present to a health facility with acute respiratory illness for the second time within 14 days. On March 21, Brunei started testing and isolating all travellers and returning residents. On March 25, SARS-CoV-2 sampling at selected sentinel health centers was introduced, followed by mandatory random screening for selected groups of foreign workers on April 7.

A confirmed case is defined as a person who tested positive for SARS-CoV-2 through real-time reverse transcriptase polymerase chain reaction (RT-PCR) test on nasopharyngeal (NP) swab (12). The first positive case in Brunei was detected on March 9, having met the testing criteria as a person with fever and cough with a recent travel history to Kuala Lumpur.

### Epidemiological investigation

Under the Infectious Disease Act, the Ministry of Health conducted epidemiological investigation and data collection for each case and close contact using WHO’s First Few Cases protocol (13). The first case was interviewed for demographic characteristics, clinical symptoms, travel history, activity mapping, and contact history. Upon identification of his participation at the Tablighi event in Malaysia, and we found that a number of other Bruneians had also participated at the same event. We subsequently obtained the details of all Bruneian participants.

NP swabs were collected from all identified participants and tested with RT-PCR. Those who tested positive were admitted to the National Isolation Centre (NIC), while those tested negative were quarantined for 14 days from their return to the country at a designated community quarantine facility where symptom and temperature screening was conducted daily. Those who developed symptoms were re-tested. Activity mapping of confirmed cases was conducted, and contact tracing initiated.

A close contact is defined as any person living in the same household, or someone within one meter of a confirmed case in an enclosed space for more than 15 minutes. All close contacts of confirmed cases were tested with RT-PCR. Those who tested positive were admitted to NIC, while those tested negative, were placed under home quarantine for 14 days from last exposure to the confirmed case. For individuals under home quarantine, their compliance and health status were monitored daily, through video calls or face-to-face assessments. Those who developed symptoms during home quarantine were re-tested.

### Clinical management

All confirmed cases were treated and isolated at NIC and followed-up until recovery. We obtained clinical information on their history (including any prior presentation to health services), examination, laboratory and radiological results from digital inpatient records on the national health information system database. Additionally, oral history was taken to ascertain whether they had symptoms up to 14 days prior to diagnosis. Cases were discharged following two consecutive negative specimens collected at ≥ 24-hour intervals.

### Cases

We categorised cases into two groups: primary (those presumably infected at the Tablighi event in Malaysia), and non-primary (those who did not attend the Tablighi event but had an epidemiological link to the primary cases).

For each case, the symptom status was recorded and classified as: (i) symptomatic, if symptoms were reported during or prior to NP swab collection; (ii) presymptomatic, if symptoms were reported after NP sampling but during admission; or (iii) asymptomatic, if no symptoms were ever reported up to discharge.

### Close contacts

We classified the type of close contact into five settings: household, relatives, workplace, social, and local religious gathering.

‘Household’ is defined as those living in the same household. Among the household members, their relationship with a case was also recorded and classified as spouse, child, and others (including other familial relationship or housekeepers living in the same household). ‘Relatives’ is defined as those who live outside the household but are related to the case. ‘Workplace’ is defined as contacts encountered in the workplace or school. ‘Social’ is defined as those encountered during travel or in social events. Lastly, ‘local religious gathering’ is defined as those who attended a local religious event in Brunei on March 5, which ran throughout the night with participants staying overnight.

### Data analysis

Group comparison was done between primary and non-primary cases using Chi-square, Fisher’s Exact, or Mann-Whitney’s tests as appropriate. Incubation period was calculated from cases where dates of exposure and symptom onset were clear. Serial interval (SI) was calculated by subtracting the date of symptom onset of the infectee from the infector; only symptomatic and pre-symptomatic infector-infectee pairs with clear epidemiological links were included.

For each setting, the attack rate (AR) was calculated by dividing the number of positive contacts by the total number of close contacts (that is, the proportion of contacts that tested positive). To identify risk factors of infection, log-binomial regression analysis was applied to estimate the risk ratio for gender, age, and setting. Further stratification was done to assess differences in infector symptom status across settings. The 95% confidence interval (95% CI) was estimated using the normal-approximation method, or binomial method if the count was less than five.

The mean observed reproductive number, R, and the distribution of personal reproductive numbers in each setting were calculated from the number of infected close contacts caused by each primary case. The 95% CI was estimated based on the Poisson distribution (14).

All analyses were conducted using Microsoft Excel and R (ver. 3.6.3) (15). A *p*-value <0.05 was considered as statistically significant. Ethical approval was obtained from the University Research Ethics Committee, Universiti Brunei Darussalam (Ref: UBD/OAVCR/UREC/Apr2020-05).

## RESULTS

### Epidemiological characteristics

Seventy-five individuals in Brunei attended the Tablighi event in Malaysia. Of these, 19 were positive for SARS-CoV-2 resulting in 52 additional cases transmitted locally, bringing the total cluster size to 71. Figure 2 illustrates the epidemiological links in the cluster by generation in the transmission chain, and by their symptomatic status. There were 32 (45.1%), 15 (21.1%), and 5 (7.0%) cases in generations one, two, and three, respectively.

**Figure 2.**
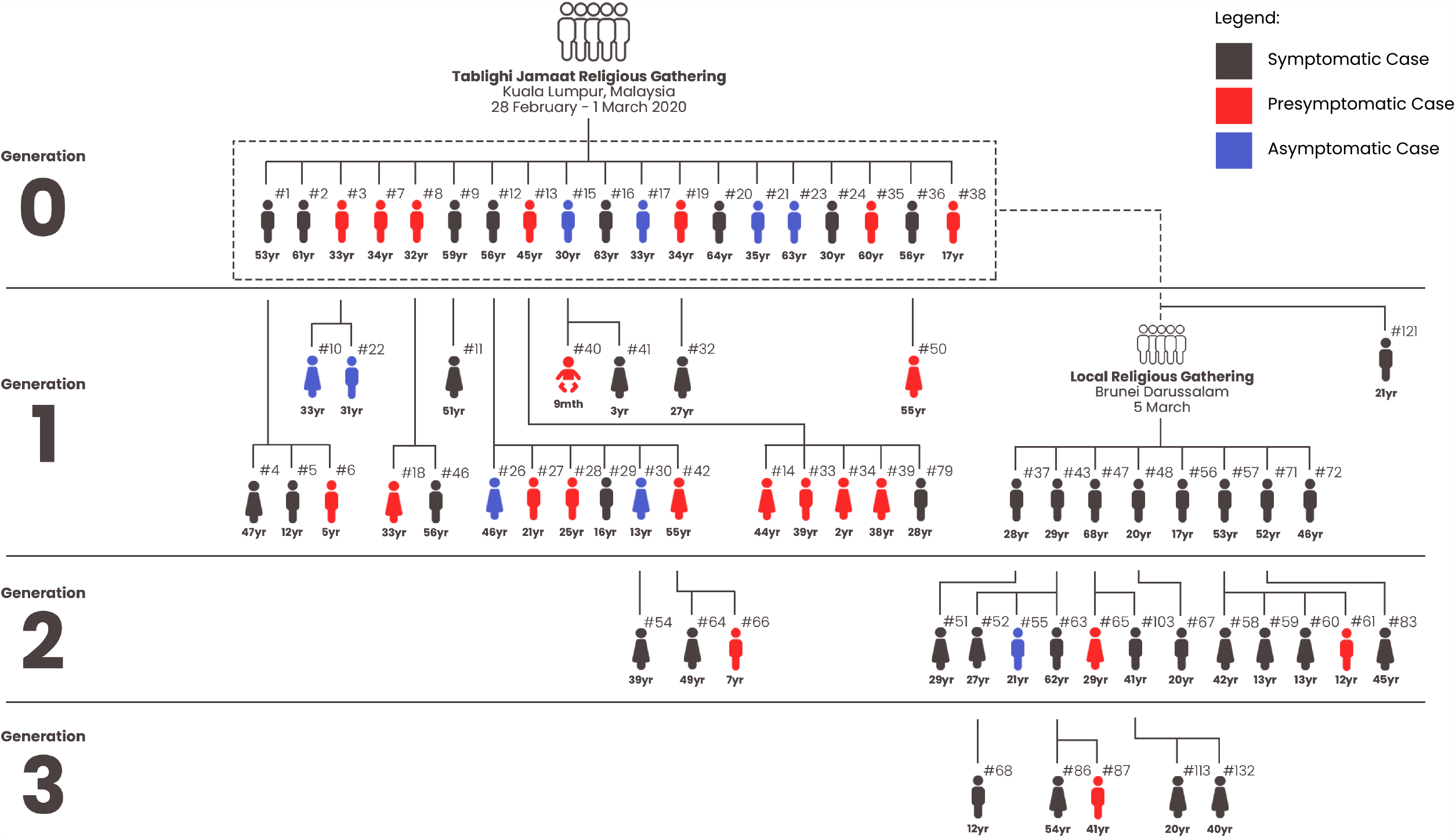
The Tablighi cluster in Brunei Darussalam, with epidemiological links illustrated by generation and symptomatic status

Table 1 shows the demographic and clinical characteristics of the cases in the cluster. The median age was 33.0 years [Interquartile range (IQR)= 29.0 years], majority male (64.8%, n=46), and 5 cases (7.1%) had pre-existing chronic conditions. Compared to non-primary cases, primary cases were significantly older and predominantly male. More than three-quarter of the cases (77.4%, n=55) were diagnosed and immediately admitted to the NIC within 5 days of symptom onset or NP swab taken (data not shown).

A substantial proportion of cases were presymptomatic (31.0%, n=22) or asymptomatic (12.7%, n=9). 40 (56.3%) patients reported symptoms during contact tracing investigation. The most commonly reported symptoms were fever, cough, and sore throat. Only one (1.4%) and two (2.8%) cases were critical and severe, respectively.

**Table 1.** Demographic and clinical characteristics of COVID-19 cases in Tablighi cluster, Brunei

The incubation period was calculated from eight cases that had confirmed epidemiological links and had attended the March 5 religious gathering in Brunei. Using their attendance date as the exposure date, the median incubation period was 4.5 days (IQR= 2.75 days, range= 1 to 11 days). Based on 35 symptomatic infector-infectee pairs, the mean (SD) SI was 4.26 (4.27) days, ranging from −4 to 17 days. Four pairs (11.4%) had negative SI values. The SI distribution resembles a normal distribution (Supplementary Figure S1).

### Transmission characteristics

Of the 1755 close contacts in the Tablighi cluster, 51 local transmissions were detected, giving an overall non-primary AR of 2.9% (95% CI: 2.2, 3.8). Case 121 (see Figure 2) was excluded from this analysis because he was not detected during contact tracing. Highest AR was observed among spouse (41.9% [95% CI: 24.1, 60.7]), followed by the local religious gathering (14.8% [95% CI: 7.1, 27.7]) and children (14.1% [95% CI: 7.8, 23.8]). The overall household AR is 10.6% (95% CI: 7.3, 15.1).

Multiple log-binomial regression analyses revealed that type of close contact was the only statistically significant variable (*p*<0.001; Table 2). When compared to social contacts, spouses of positive cases had the highest adjusted risk ratio of getting the infection (45.20 [95% CI: 16.8, 156.1]), followed by the local religious gathering attendees (15.60 [95% CI: 4.8, 59.9]), and their children (14.09 [95% CI: 4.8, 51.5]).

**Table 2.** Risk factors of SARS-COV2 infection among close contacts

ARs also differed by symptom status of the infector (Table 3). ARs in households where the infectors were symptomatic (14.4%) were higher than those who were asymptomatic (4.4%) or presymptomatic (6.1%). AR for the local religious gathering could not be calculated as three primary cases at the event had different symptom status; hence we could not ascertain how transmission occurred. In the household setting, symptomatic cases have 2.66 times higher risk of transmitting to their close contacts, when compared to asymptomatic and presymptomatic (crude risk ratio: 2.66 [95% CI: 1.12, 6.34], Table S1).

**Table 3.** Attack rates in different settings, stratified by symptom status of the infector

The mean observed R was highest in the local religious gathering (2.67), followed by the household setting (0.67 [95% CI: 0.44, 0.96]) (Table 4). The distribution of the observed R in the household setting was skewed towards zero (Figure S2). 71.4% of household infections (20 of 28 positive contacts) were from 16.7% of cases (7 of 42 possible links).

**Table 4.** Characteristics and mean observed R for each setting

## DISCUSSION

We characterise the Tablighi Jama’at cluster that started the COVID-19 epidemic in Brunei. Our analysis reveals several key findings. First, SSE plays an important role in SARS-CoV-2 transmission. Second, there is high transmission variability across different settings. Third, transmission varies between symptomatic vs. asymptomatic and presymptomatic cases, and we highlight the potential for silent chains of transmission.

### SSE

Within this cluster, 38% of all cases were participants at an SSE: 19 (26.7%) from the Tablighi event in Malaysia, and eight (11.3%) from the local religious gathering in Brunei. Notably, 19 of the 75 Brunei attendees at the Tablighi event tested positive. Assuming a representative sample, this would suggest an AR of 25%, implying that approximately 4,000 cases (of an estimated 16,000 participants) may have been infected at that Malaysian event. Moreover, we find that the highest overall non-primary AR and mean observed R was at the local religious gathering (AR= 14.8%, R= 2.67), which was higher than that observed in the household setting (AR= 10.6%, R= 0.67). These observations suggest a role for mass gatherings in facilitating SARS-CoV2 transmission.

The Tablighi is an apolitical Islamic movement with a presence in nearly 200 countries. Tablighi adherents usually travel to gather at annual international events each lasting several days. Communal prayers, meals and speeches form part of these events. In Malaysia, the participants stayed and slept at the mosque, and several participants were deputised to cook meals and clean. Smaller gatherings, usually every week or so take place in their home countries, with other adherents. Over the course of this investigation, we identified several common characteristics at both the local religious gathering and the Tablighi event in Malaysia (11). First, significant numbers of people gathered in an enclosed area for a prolonged period. Second, attendees had a recent travel history – the Tablighi event in Malaysia drew participants from across the world, while the local religious gathering had at least three individuals who had recently returned from Malaysia. Third, communal sharing of sleeping areas and toilets, and shared dining were observed. We propose that these three characteristics are hallmarks for the development of SSE for SARS-CoV-2 transmission and can be used as ‘red flags’ by health authorities in their risk assessment and mitigation strategies for preventing and detecting high risk activities including mass gatherings, and other institutional settings such as care homes, prisons and dormitories.

### Variability of non-primary AR across different settings

To a lesser degree, our observations on the within-household transmission are similar to that observed for the two religious gatherings. Out of 16 household contacts who subsequently became first generation cases, 10 (62.5%) of these were from just three primary cases. As such, even within similar settings, we can expect wide variability in transmission patterns. This observation supports our finding of a moderately high household AR but an observed R of less than one, suggesting that transmission is driven by a relatively small number of cases (5). High ARs in spouses and children reflect intimate relationships with high degree of interaction, close proximity, and in the case of the spouse, sleeping in the same room. Concordant with our SSE findings, we suggest that encounters among groups of people that involve close proximity in enclosed settings for prolonged time periods (at least for one night) is a main driver of SARS-CoV-2 transmission.

Our overall non-primary AR result of 10.6% in the household setting is comparable to other studies that used contract-tracing datasets (16-19). A study near Wuhan, China (20) reported a higher AR of 16.3%; however, they detected 56.2% of their cases more than five days after symptom onset. By contrast, 77.4% of the cases in our study were detected and isolated within five days of symptom onset, suggesting that early case isolation can reduce AR. The aggressive testing of contacts strategy employed may have contributed to this.

We note the low non-primary AR (<1%) and mean observed R (<0.3) for workplace and social settings. While moderate physical distancing was implemented in Brunei following the identification of this cluster, there was no community quarantine or lockdown, public services and businesses remained open, and no internal movement restrictions were imposed.

Combined with our observations on the role of SSE in driving SARS-CoV-2 transmission, we suggest that in areas with limited community transmission full lockdown measures that adopt a blunt approach by restricting all movement can be avoided, in favour of a more targeted approach that includes a combination of case isolation, contact tracing, and moderate levels of physical distancing that take into account the ‘red flags’ for mass gatherings identified earlier. However, this approach is resource intensive and only feasible where there is sufficient public health capacity. The high asymptomatic proportion suggest that even with best efforts at contact tracing, potential for widespread community transmission is clear. Once established, suppression necessitates the implementation of broader physical distancing measures (9, 21). Nonetheless, if done effectively, contact tracing and case isolation approaches are shown to control the COVID-19 outbreak during its early stage (22). Modelling studies using South Korean data showed that less extreme physical distancing measures can help to suppress the outbreak (23).

### Symptomatic vs. asymptomatic and presymptomatic transmission

We identified several environmental (settings) and behavioural factors that potentially account for higher ARs observed in mass gatherings and in the household. In order to assess the impact of host factors in driving transmission, we compared non-primary AR in symptomatic vs. asymptomatic and presymptomatic individuals, considering the high proportion of asymptomatic (12.7%) and presymptomatic (31.0%) of cases.

While there are case reports of presumptive asymptomatic and presymptomatic transmission (24, 25), observational studies quantifying such transmission are few. A study from Ningbo, China analysed the overall ARs in symptomatic vs. asymptomatic cases and did not find significant difference between the two groups (26). Another study, re-interpreted the same data and theorized that under certain conditions, symptomatic cases could be more transmissible than asymptomatic ones (27). In fact, our overall crude risk ratio for symptomatic cases showed no significant difference when compared with asymptomatic and/or presymptomatic cases (Table 3, Table S1). However, we suggest that this masks the true picture in transmissibility when different settings are taken into account.

In our study, we do not find a significant difference in AR in non-household settings. These settings usually practice some form of non-pharmaceutical interventions (NPI)—individuals with moderate and severe symptoms may be on medical leave, and it is reasonable to expect some physical distancing would be practiced by contacts of persons who display visible symptoms. This is less feasible within the household setting; hence we suggest that transmission occurs more frequently at the household level where control measures are less practical. We observed that the household AR for symptomatic cases (14.4%) is higher than that of asymptomatic (4.1%) or presymptomatic cases (6.1%), suggesting that the presence of symptoms is a host factor in driving transmission.

The higher household AR observed among symptomatic cases suggest that testing for such cases should be prioritised, especially in low resource areas with limited testing capacity. Nonetheless, an AR of 4.4% and 6.1% in asymptomatic and presymptomatic cases, respectively, is not negligible. As such, our findings have several implications for high resource areas with greater testing capacity. First, it strengthens the argument for testing household contacts in the absence of symptoms. Second, there is a need to allow for ‘slack’ in the surveillance system as the high proportion of asymptomatic cases pose challenges for rapid detection and isolation. We recommend that even in countries with highly developed testing and tracing capacities, moderate levels of social distancing should be implemented to account for this. Third, proactive testing of travellers, attendees at ‘red flag’ events, and institutional settings may be necessary to contain COVID-19 spread.

This study has several limitations. First, as a retrospective study based on a contact tracing dataset, the index case determination or the direction of transmission may be uncertain, particularly as a substantial proportion of cases are asymptomatic. Moreover, without accounting for outside sources of infection, setting-specific SARs could have been overestimated (although no community transmission has been detected in Brunei). Viral sequencing can confirm homology between the strains infecting the index and secondary cases across the various settings, however this was not conducted for all cases. Second, we have not accounted for other potential environmental factors such as the relative size of the household, time spent at home with others, air ventilation, and indirect transmission through fomites. Third, we do not have information on NPIs practiced by the close contacts; presumably, individuals would take precautions during an outbreak. Fourth, symptom status of the cases was reported during their swab collection date. We assume this to be reflective of their actual condition when their close contacts were exposed, however, this may not be necessarily true for all cases. Finally, the generalizability of our results are limited due to no community transmission, a lack of cases in settings such as residential care facilities and dormitories, and small number of cases.

The main strength of our study is the availability of a complete contact tracing dataset at the national level. Since all contacts were tested, it is reasonable to assume that this study more accurately detects SARS-CoV-2 transmission than those that only test symptomatic contacts. In conclusion, our analysis highlights the variability of SARS-CoV-2 transmission across different settings and in particular, the role of SSEs. We identify ‘red flags’ for the development of potential SSEs, and describe environmental, behavioural, and host factors that drive transmission. Overall, we provide evidence that a combination of case isolation, contact tracing, and moderate physical distancing measures is an effective approach to containment.

## Data Availability

De-identified participant data is available upon request to the corresponding author.

## Acknowledgement

The authors would like to thank Mr Haji Mohamad Ruzaimi Haji Rosli from Corporate Communications, Ministry of Health Brunei, for his assistance in designing the figure for this manuscript.

## Supplementary information

**Figure S1**. Distribution of the serial interval, fitted with a normal distribution

**Figure S2**. Distribution of the household observed R

Table S1. Attack rates in different settings, stratified by symptom status of the infector (symptomatic versus presymptomatic and asymptomatic)

## References

1. Ong SWX, Tan YK, Chia PY, Lee TH, Ng OT, Wong MSY, et al. Air, Surface Environmental, and Personal Protective Equipment Contamination by Severe Acute Respiratory Syndrome Coronavirus 2 (SARS-CoV-2) From a Symptomatic Patient. JAMA. 2020.

2. Furukawa N, Brooks J, Sobel J. Evidence Supporting Transmission of Severe Acute Respiratory Syndrome Coronavirus 2 While Presymptomatic or Asymptomatic. Emerging Infectious Disease journal. 2020;26(7).

3. He X, Lau EHY, Wu P, Deng X, Wang J, Hao X, et al. Temporal dynamics in viral shedding and transmissibility of COVID-19. Nature Medicine. 2020 2020/04/15.

4. Kucharski AJ, Russell TW, Diamond C, Liu Y, Edmunds J, Funk S, et al. Early dynamics of transmission and control of COVID-19: a mathematical modelling study. The Lancet Infectious Diseases. 2020.

5. Liu Y, Eggo RM, Kucharski AJ. Secondary attack rate and superspreading events for SARS-CoV-2. The Lancet. 2020;395(10227):e47.

6. Stein ML, van der Heijden Pgm, Buskens V, van Steenbergen JE, Bengtsson L, Koppeschaar CE, et al. Tracking social contact networks with online respondent-driven detection: who recruits whom? BMC Infectious Diseases. 2015 2015//;15(1):1–12.

7. Li Y, Yu ITS, Xu P, Lee JHW, Wong TW, Ooi PL, et al. Predicting Super Spreading Events during the 2003 Severe Acute Respiratory Syndrome Epidemics in Hong Kong and Singapore. Am J Epidemiol. 2004;160(8):719–28.

8. !!! INVALID CITATION !!! [Frieden, 2020 #4083].

9. Lau H, Khosrawipour V, Kocbach P, Mikolajczyk A, Schubert J, Bania J, et al. The positive impact of lockdown in Wuhan on containing the COVID-19 outbreak in China. Journal of Travel Medicine. 2020.

10. Department of Economic Planning and Development. Mid-year population estimates for Brunei Darussalam, 2019. 2020 [cited 2020 22 April]; Available from: http://www.deps.gov.bn/SitePages/Population.aspx

11. Mat NFC, Edinur HA, Razab Mkaa, Safuan S. A Single Mass Gathering Resulted in Massive Transmission of COVID-19 Infections in Malaysia with Further International Spread. Journal of Travel Medicine. 2020.

12. Wong J, Koh WC, Momin RN, Alikhan MF, Fadillah N, Naing L. Probable causes and risk factors for positive SARS-CoV-2 test in recovered patients: Evidence from Brunei Darussalam. Journal of Medical Virology. 2020 2020/06/19;n/a(/a).

13. World Health Organization. The First Few X (FFX) Cases and contact investigation protocol for 2019-novel coronavirus (2019-nCoV) infection, version 2. 2020.

14. Liu Y, Gayle AA, Wilder-Smith A, Rocklöv J. The reproductive number of COVID-19 is higher compared to SARS coronavirus. Journal of Travel Medicine. 2020;27(2).

15. R Core Team. R: A language and environment for statistical computing. R Foundation for Statistical Computing, Vienna, Austria; 2020.

16. Cheng H-Y, Jian S-W, Liu D-P, Ng T-C, Huang W-T, Lin H-H. High transmissibility of COVID-19 near symptom onset. medRxiv. 2020:2020.03.18.20034561.

17. Luo L, Liu D, Liao X-l, Wu X-b, Jing Q-l, Zheng J-z, et al. Modes of contact and risk of transmission in COVID-19 among close contacts. medRxiv. 2020:2020.03.24.20042606.

18. Bi Q, Wu Y, Mei S, Ye C, Zou X, Zhang Z, et al. Epidemiology and Transmission of COVID-19 in Shenzhen China: Analysis of 391 cases and 1,286 of their close contacts. medRxiv. 2020:2020.03.03.20028423.

19. Jing Q-L, Liu M-J, Yuan J, Zhang Z-B, Zhang A-R, Dean NE, et al. Household Secondary Attack Rate of COVID-19 and Associated Determinants. medRxiv. 2020:2020.04.11.20056010.

20. Li W, Zhang B, Lu J, Liu S, Chang Z, Cao P, et al. The characteristics of household transmission of COVID-19. Clinical Infectious Diseases. 2020.

21. Sjödin H, Wilder-Smith A, Osman S, Farooq Z, Rocklöv J. Only strict quarantine measures can curb the coronavirus disease (COVID-19) outbreak in Italy, 2020. Eurosurveillance. 2020;25(13):2000280.

22. Hellewell J, Abbott S, Gimma A, Bosse NI, Jarvis CI, Russell TW, et al. Feasibility of controlling COVID-19 outbreaks by isolation of cases and contacts. The Lancet Global Health. 2020.

23. Park SW, Sun K, Viboud C, Grenfell BT, Dushoff J. Potential roles of social distancing in mitigating the spread of coronavirus disease 2019 (COVID-19) in South Korea. medRxiv. 2020:2020.03.27.20045815.

24. Wei WE, Li Z, Chiew CJ, Yong SE, Toh MP, Lee VJ. Presymptomatic Transmission of SARS-CoV-2 — Singapore, January 23–March 16, 2020. MMWR Morb Mortal Wkly Rep. 2020.

25. Qian G, Yang N, Ma AHY, Wang L, Li G, Chen X, et al. COVID-19 Transmission Within a Family Cluster by Presymptomatic Carriers in China. Clinical Infectious Diseases. 2020.

26. Chen Y, Wang A, Yi B, Ding K, Wang H, Wang J, et al. The epidemiological characteristics of infection in close contacts of COVID-19 in Ningbo city Chinese Journal of Epidemiology. 2020;41.

27. He D, Zhao S, Lin Q, Zhuang Z, Cao P, Wang MH, et al. The relative transmissibility of asymptomatic cases among close contacts. International Journal of Infectious Diseases. 2020;94:145–7.

